# COVID-19 Control Strategies and Intervention Effects in Resource Limited Settings: A Modeling Study

**DOI:** 10.1101/2020.04.26.20079673

**Authors:** Kiran Raj Pandey, Anup Subedee, Bishesh Khanal, Bhagawan Koirala

**Author notes:** <u>Corresponding Author</u> Kiran Raj Pandey, Hospital for Advanced Medicine and Surgery, Budhanilkantha-1, Kathmandu, 44600, Nepal.

## Abstract

**Background:** Many countries with weaker health systems are struggling to put together a coherent strategy against the COVID-19 epidemic. We explored COVID-19 control strategies that could offer the greatest benefit in resource limited settings.

**Methods:** Using an age-structured SEIR model, we explored the effects of COVID-19 control interventions--a lockdown, physical distancing measures, and active case finding (testing and isolation, contact tracing and quarantine)-- implemented individually and in combination to control a hypothetical COVID-19 epidemic in Kathmandu (population 2.6 million), Nepal.

**Results:** A month-long lockdown that is currently in place in Nepal will delay peak demand for hospital beds by 36 days, as compared to a base scenario of no interventions (peak demand at 108 days (Inter-Quartile Range IQR 97–119); a 2 month long lockdown will delay it by 74 days, without any difference in annual mortality, or healthcare demand volume. Year-long physical distancing measures will reduce peak demand to 36% (IQR 23%-46%) and annual morality to 67% (IQR 48%-77%) of base scenario. Following a month long lockdown with ongoing physical distancing measures and an active case finding intervention that detects 5% of the daily infection burden could reduce projected morality and peak demand by more than 99%.

**Interpretation:** Limited resources settings are best served by a combination of early and aggressive case finding with ongoing physical distancing measures to control the COVID-19 epidemic. A lockdown may be helpful until combination interventions can be put in place but is unlikely to reduce annual mortality or healthcare demand.

## Introduction

The COVID-19 epidemic, first reported in China in December 2019, has now spread to more than 200 countries and has been declared a global pandemic by the World Health Organization (WHO). ^1^ In the past three months, many countries have struggled to put together a coherent strategy against the epidemic, even while more than two million infections and 160,000 deaths have been reported.^2^ Countries with limited resources are likely to face even greater difficulties against this epidemic, as they grapple with limited resources and fewer intervention options.^3–5^

Control strategies and interventions against the spread of COVID-19 center around three broad options: one, reduce the number of infectious individuals; two, reduce the number of susceptible individuals; and three, reduce contact between susceptible and infectious individuals.^6–9^ In the absence of a vaccine against the COVID-19 causing SARS-CoV2 virus, or an effective treatment, available control strategies are limited. They include ones that aim to reduce the burden of infectious individuals (by identifying and isolating infectious individuals) and ones that aim to reduce contact (with physical distancing measures, or a lockdown). Countries like Taiwan and Vietnam have been able to contain the epidemic by means of meticulous public health measures including aggressive testing, isolation, contact tracing and quarantine combined with border entry monitoring.^10–12^ Korea has similarly been able to mitigate the epidemic by means of aggressive testing and isolation combined with physical distancing measures.^13,14^ China on the other hand has managed to suppress the epidemic by means of a strategy involving strictly enforced lockdowns, aggressive testing isolation and quarantine. ^15,16^

Countries like Nepal, which have been declared a high risk country for a COVID-19 epidemic by the WHO, stand to face difficult strategic choices and challenges given the limited amount of healthcare resources. These challenges have been evident in the last three months. On January 24, 2020 Nepal became the first country in South Asia to report a SARS-CoV2 infection--in a student who had returned from Wuhan, China. Although no new cases were reported until March 23 (since then, as of April 20, 29 more cases have been reported),^17^ Nepal tried to put together a response against a possible epidemic: First, as a precautionary measure the government began limiting international air travel from affected countries in February and later from the rest of the world. Land border crossings with India remained open until they were finally closed on March 24, 2020. ^18^

Pre-emptive physical distancing measures were introduced in the middle of March, when schools were closed and annual exams canceled. The public was advised to avoid all non-essential events and gatherings. Long distance buses were closed on March 23, 2020 following which a nationwide lockdown--which has now been extended until April 27--was declared on March 24. In these three months less than 10 000 RT-PCR based tests (<3 per 10 000 people) for SARS-CoV2 have been carried out in the country.^17^ Testing volumes have been low due to minimal testing kit and reagent availability, strict testing criteria, as well as limited testing locations.

Even while Nepal has made attempts to put off a potential epidemic in the country, its response has been hindered by a lack of clarity on what the best strategy on stopping a potential epidemic might be. To study this, we built a mathematical model to simulate the consequences of adopting various strategies in preventing or controlling a possible COVID-19 epidemic in Kathmandu, Nepal. We first estimated the burden of an unmitigated epidemic. We then assessed the potential effects of implementing control interventions: a lockdown (total shutdown of movement), physical distancing (avoiding large gatherings, no handshake, minimizing physical proximity), or aggressive testing and contact tracing with quarantine. In addition, we explored the effect of these interventions when implemented together. We analysed potential demand for health services and the relative mortality burden in each of these scenarios.

## Methods

We used an age-structured SEIR (Susceptible, Exposed, Infectious, Recovered) model with heterogeneous mixing to estimate epidemic burden due to COVID-19 in Kathmandu.^19,20^ We created additional compartments Q, J, H, and U for individuals in quarantine, isolation, hospital and ICU respectively. Compartment D represents individuals that are dead. These compartments were further divided into 16 age-structured groups each and populated based on the age-specific population distribution of Nepal. Population mixing patterns were given by a Nepali population specific contact matrix.^21^

In the model, susceptible individuals in the age-group *i* move from the compartment (*S_ij_*) to the exposed compartment (*E_ij_*) based on the force of infection (λ_*ij*_). The force of infection is obtained by multiplying the transmission rate β*_ij_* to the proportion of infectious contacts. β_*ij*_ is the transmission rate when a susceptible person in age group *i* mixes with an infectious person in age group *j*. The age-dependent transmission rate is expressed as follows:

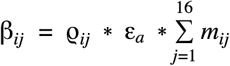

where the probability of infection per contact (ϱ*_ij_*) has a Poisson distribution given by,

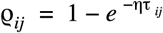

And, the force of infection is,

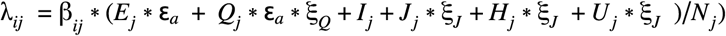

Contact rates for individuals in group *i* with group *j* are given by the following contact matrix,

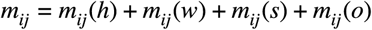

Where (h) home, (w): work, (s): school and (o): other place of contact. Interventions that change contact patterns give a new contact matrix that reflects the change (figure 1).

**Figure 1.**
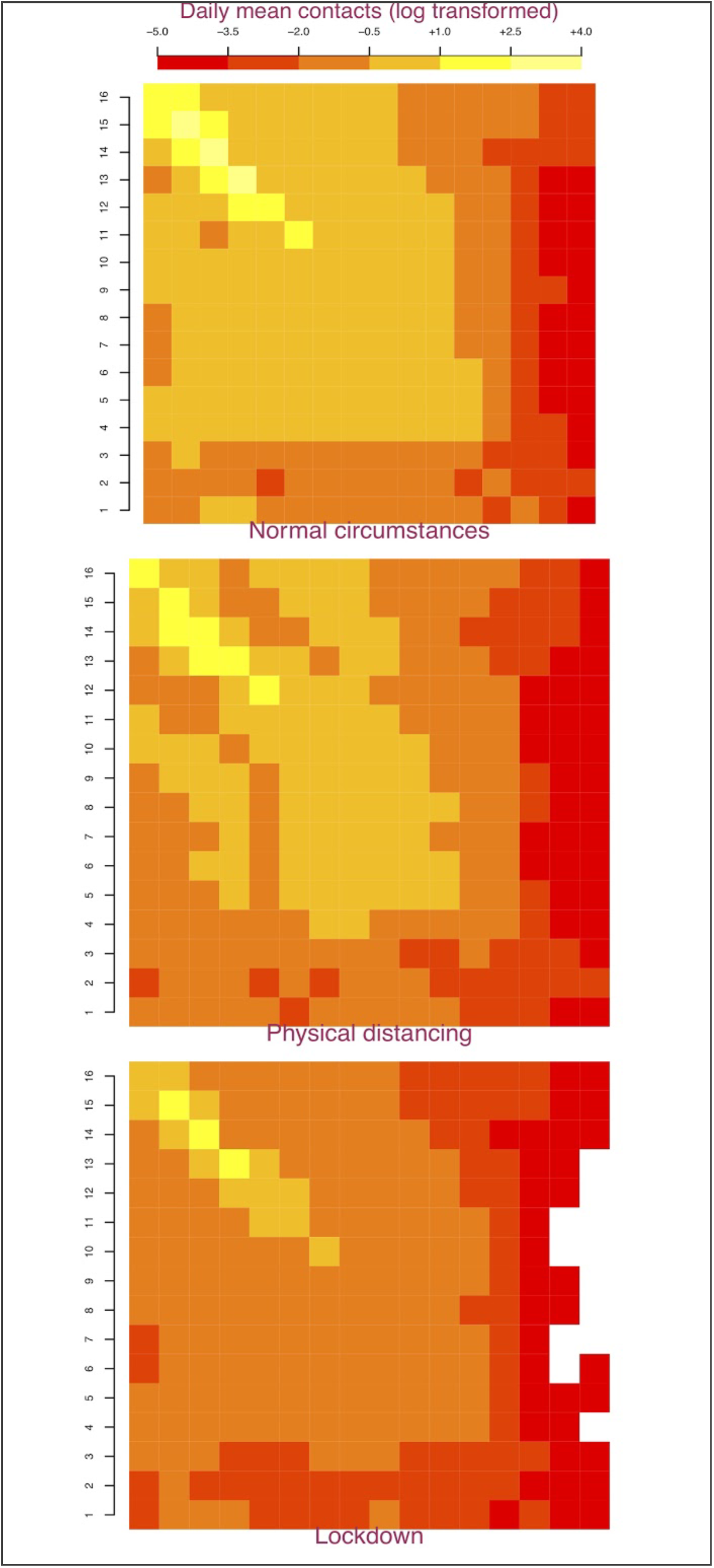
Age and intervention specific contact matrices for Nepal. In these matrices, the population is divided into 16 5-year age cohorts represented along the two axes. The top left square represents interactions between 0–4 year olds and the bottom right aquare represents contacts between individuals who are 75 years and older. Colors in the squares represent the log transformed mean daily contact rate between corresponding 5-year age cohorts. Dark/red colors represent fewer contacts, light/yellow colors represent a greater number of contacts. Physical distancing causes the lighter squares to darken indicating fewer contacts, while a lockdown causes the lighter squares to darken even further.

ρ**_ij_** is the probability of transmission per contact between an individual in group *i* with *j*. η is the number of infections transmitted per unit time and τ_ij_ is the duration of each contact between individuals in groups i and j. Exposed individuals move to infectious compartment I at a rate **σ** (incubation rate), where 1/**σ** is the incubation period, and to quarantine at rate **ϕ_1_**. People who have been exposed but are asymptomatic are infectious but with a reduced proportion given by ε_a_. We assume ε_a_ of 0·10 for our analysis.^22^ Similarly infectious people move to isolation J at a rate ϕ**_2_**.*ξ_J_* and *ξ_Q_* are infectiousness factors for individuals in isolation and quarantine respectively, which we estimate at 0·15 and 0·30 based on the extent to which their contact with susceptible individuals is reduced.^21^ For our analysis we assume that individuals sick enough to require hospitalization (compartments H and U) will either be isolated at the hospital, or self-isolate at home if no hospital beds are available.

We assume all deaths happen only among individuals who require hospitalization. The general ward mortality ratio (g.fr_i_) is (ϰ*_i_* /h.r_i_)^*^(1-f) and ICU mortality ratio, represented by u.fr_i_ is (ϰ*_i_* /h.r_i_)^*^(f), where ϰ*_i_* is infection fatality ratio for the ith age-group, h.r_i_ is the proportion of individuals in age-group *i* who require hospitalization. We assume that, of all the deaths, a fraction f (which we assume to be 0·8) of deaths occur among individuals who require ICU. γ is the recovery rate among infectious individuals that don’t require hospitalization, δ_1_ is the recovery rate among general ward patients and δ_2_ is the recovery rate among patients who require ICU. Additionally, fatality ratio increases by a factor *hs.f* when health services are overburdened. Its calculation is explained further below.

Our model is described by the flow diagram in figure 2, which corresponds to the following system of differential equations, where 1 <= i <= 16 and represents 16 5-year age-cohorts.

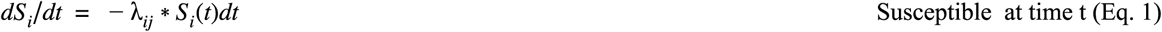

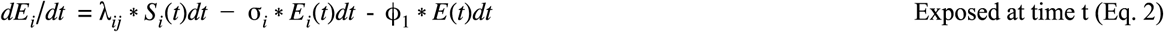

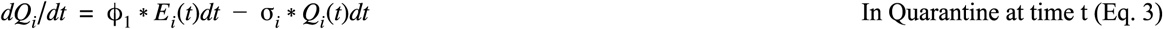

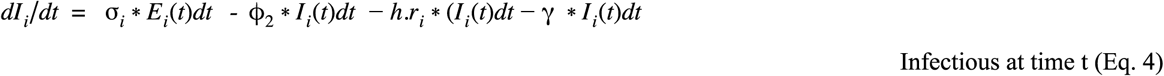

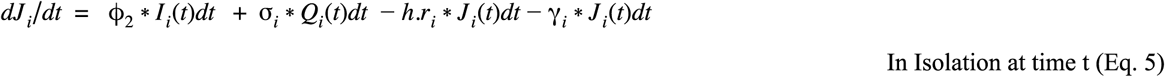

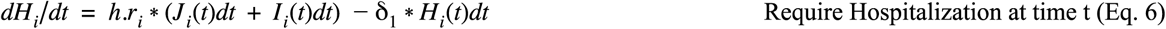

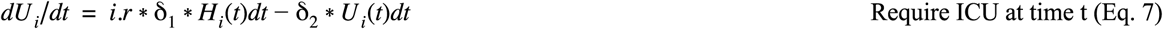

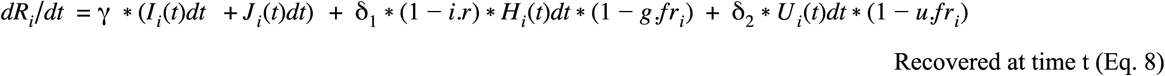

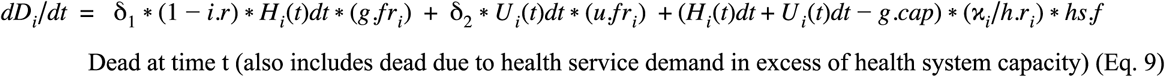

**Figure 2.**
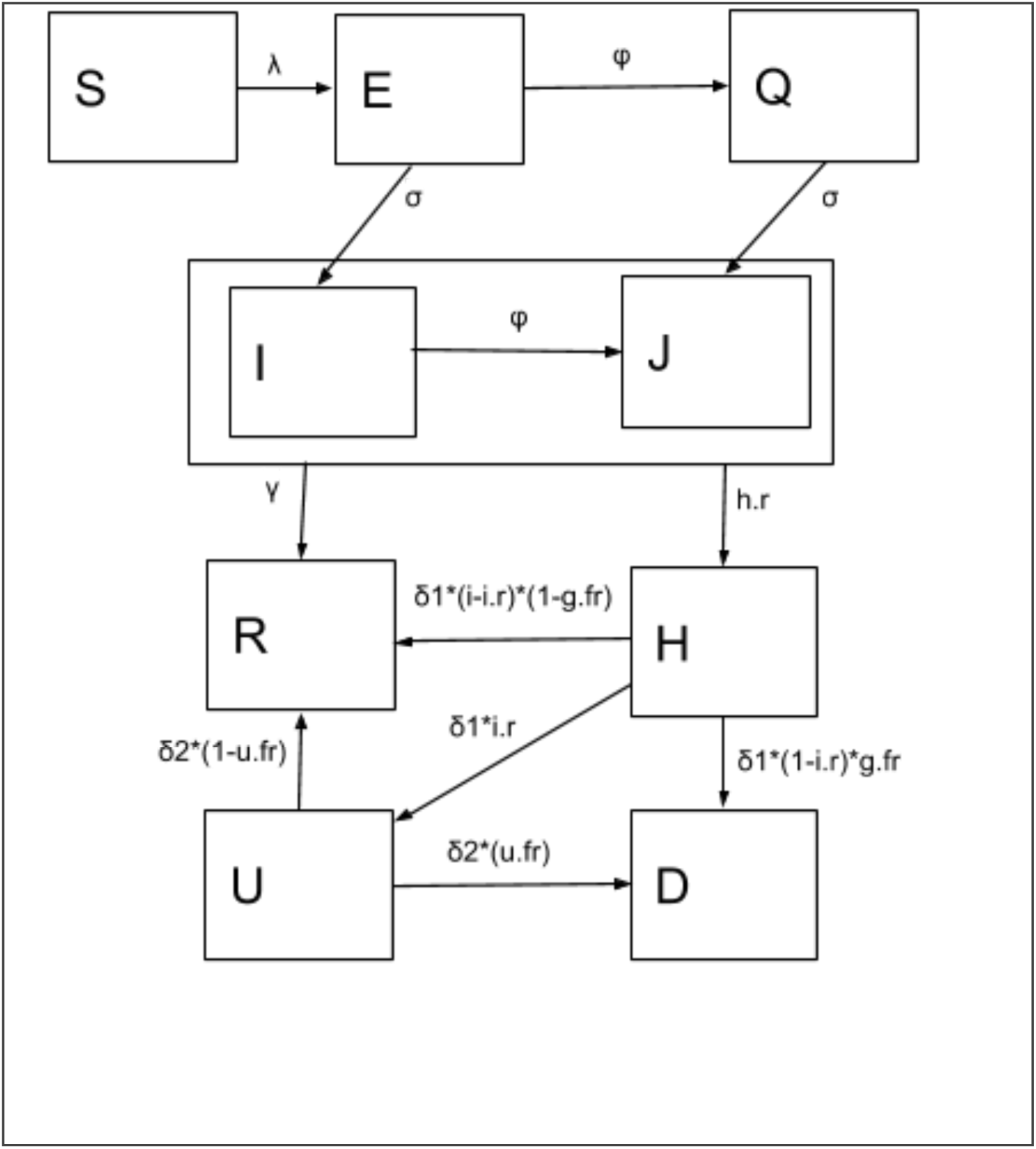
Flow diagram of the SEIR model for COVID-19 transmission dynamics. Boxes represent disease state compartments for Susceptible (S), Exposed (E), Infectious (I), and Removed (R), with additional compartments for Quarantine (Q), Isolation (J), Hospitalized in the general ward (H), Hospitalized in the ICU (U) and Dead (D). Arrows represent flow between compartments with the flow determined by their corresponding parameter values. Parameter values are further explained in table 1.

This system of 9 equations then gives us 16 equations for each age-group resulting in a total of 144 equations for the simulation. These ordinary differential equations are solved using a solver in the deSolve package in R (v 3·6·3).

## Model Parameters

The model was parameterized using published estimates of COVID-19 epidemic dynamics. We first sampled the Ro from a uniform distribution in the interval [2, 2·8] and used it to calculate the transmission rate based on age-specific contact rates.^23,24^ We sampled the incubation period from a uniform distribution in the interval [3, 7] days; we used an infectious duration of 7 days.^23,25^ We obtained age-specific hospitalization rates and mortality ratios among infected individuals from a recent analysis from China.^26^ 20% of those who require hospitalization are assumed to require ICU care. ^27,28^ Recovery among those who require general ward hospitalization occurs at rate δ_1_ and the ICU at rate δ_2_ and are calculated as the reciprocal of their respective lengths of stay. Infectious people who do not require hospital admission recover at a rate γ. We used published estimates of age-specific hospitalization rates for infected individuals to calculate the number of people who need hospitalization.^26^ Adjusting these estimates from China to the Nepali population gives an age-adjusted hospitalization rate of 2·8%. We assume that all patients hospitalized in the general ward require an eight day hospital stay on average; ICU stays are six days on average. Total hospital bed capacity is estimated at 5400 (including 250 ICU beds).^28^ Parameter values are given in table 1.

**Table 1:**
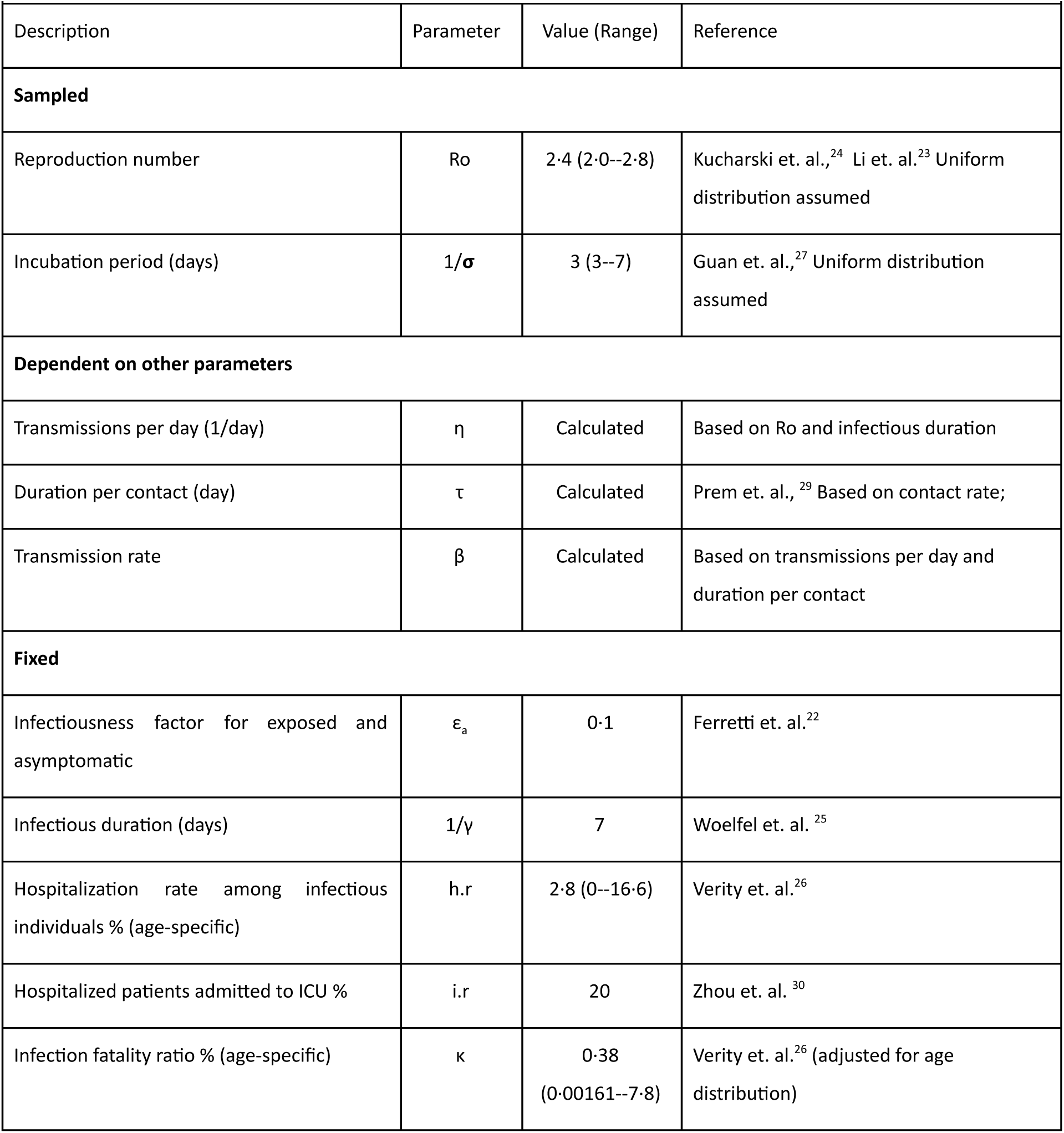
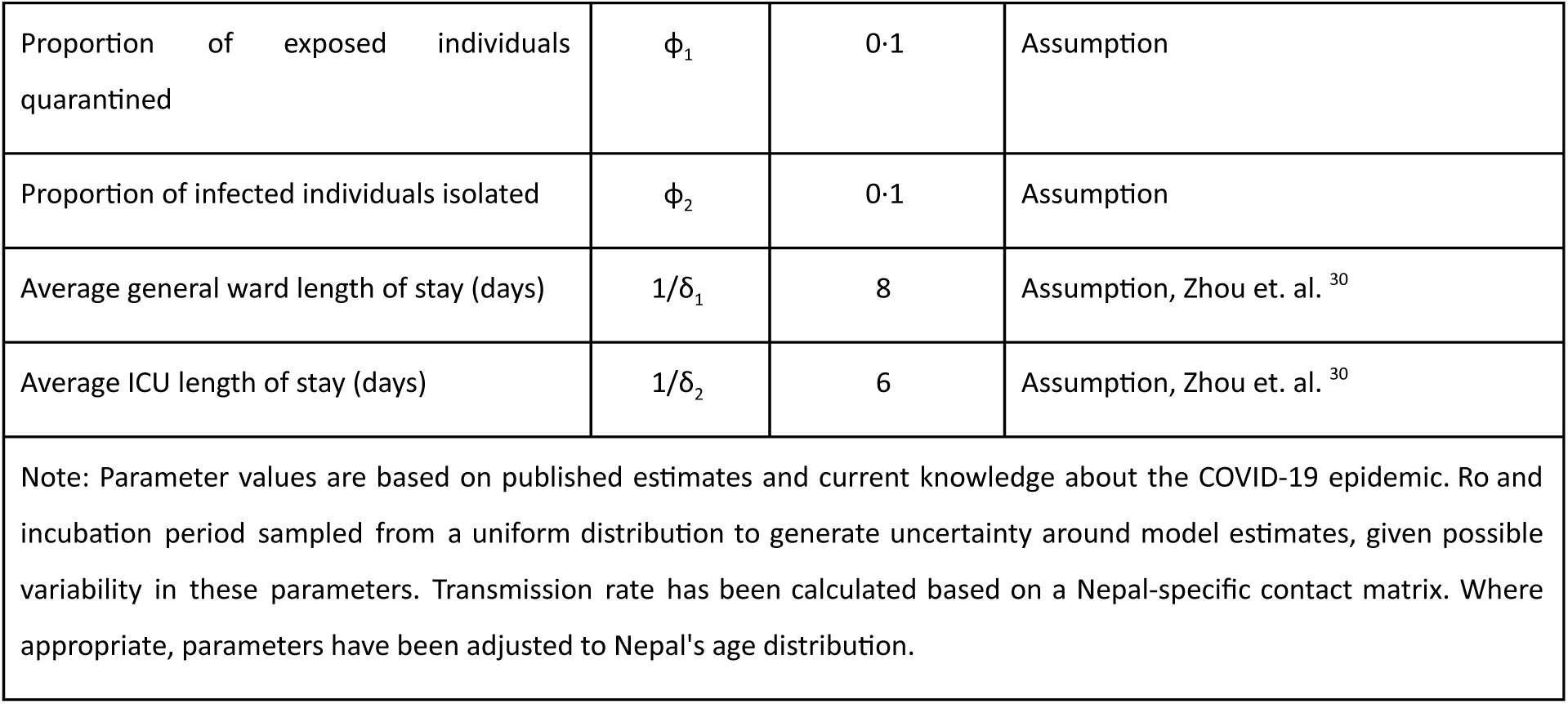
Parameter values for COVID-19 transmission dynamics in Kathmandu, Nepal

## Interventions

A total lockdown is assumed to reduce overall contacts by 70%. Physical distancing is estimated to be only 50% as effective as lockdown in reducing contacts. Individuals placed in isolation are assumed to have contacts reduced by 75% at home and 90% across all other settings. Individuals placed in home quarantine have the same reductions but with only 50% compliance.^31,32^ These estimates are summarized in Table 2. Since a lockdown is already in place, we did not analyze the individual effect of school and workplace closures. Multi-intervention epidemic control strategies were created by a linear combination of these individual strategies. We also used time-dependent control variables to simulate the effect of time varying interventions (figure 3).

**Table 2:**
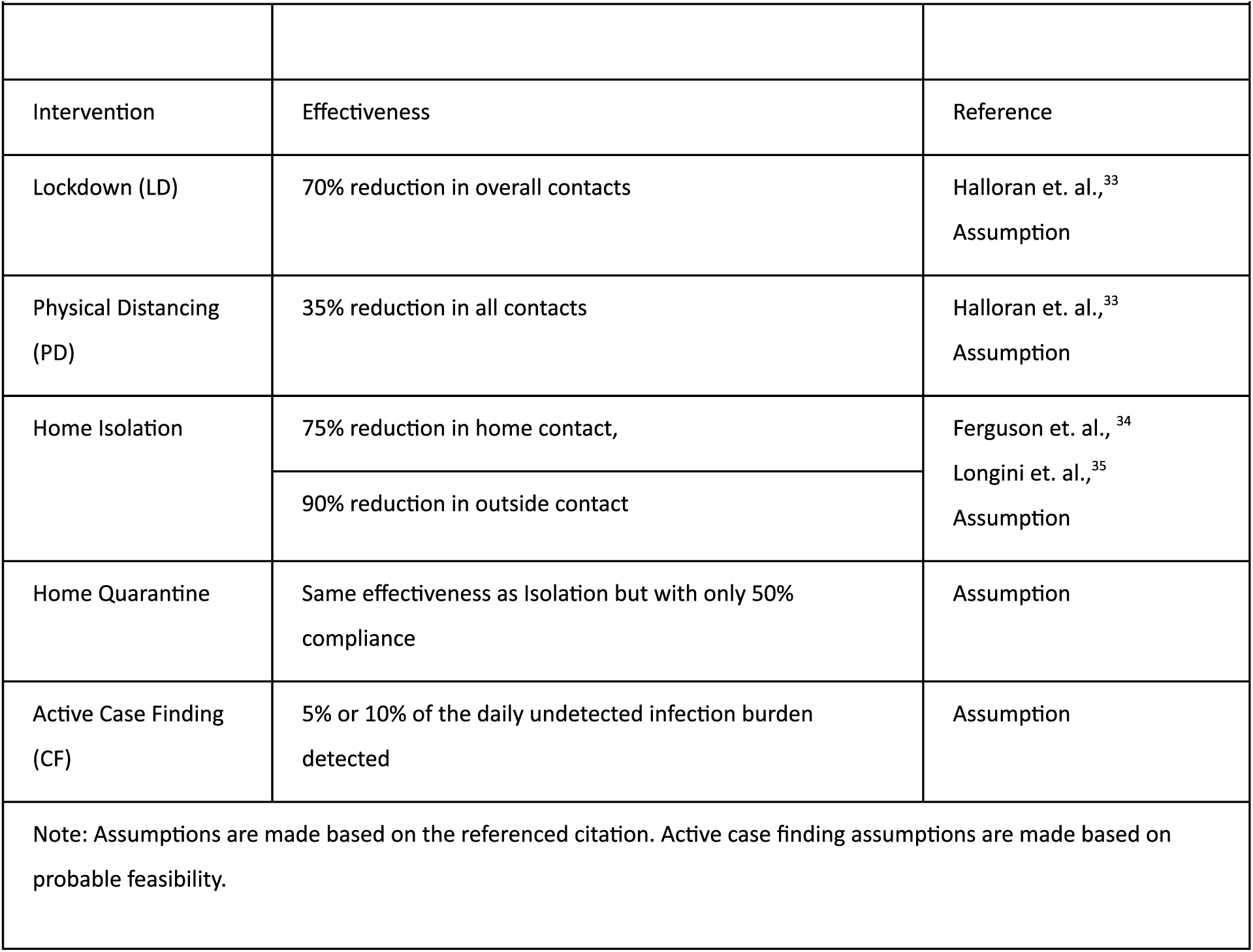
Interventions against COVID-19 and their effectiveness

| Table 2: Interventions against COVID-19 and their effectiveness |  |  |
| --- | --- | --- |
| Intervention | Effectiveness | Reference |
| Lockdown (LD) | 70% reduction in overall contacts | Halloran et. al.,<sup>33</sup><br>Assumption |
| Physical Distancing (PD) | 35% reduction in all contacts | Halloran et. al.,<sup>33</sup><br>Assumption |
| Home Isolation | 75% reduction in home contact, | Ferguson et. al.,<sup>34</sup><br>Longini et. al.,<sup>35</sup><br>Assumption |
|  | 90% reduction in outside contact |  |
| Home Quarantine | Same effectiveness as Isolation but with only 50% compliance | Assumption |
| Active Case Finding (CF) | 5% or 10% of the daily undetected infection burden detected | Assumption |
| Note: Assumptions are made based on the referenced citation. Active case finding assumptions are made based on probable feasibility. |  |  |

**Figure 3.**
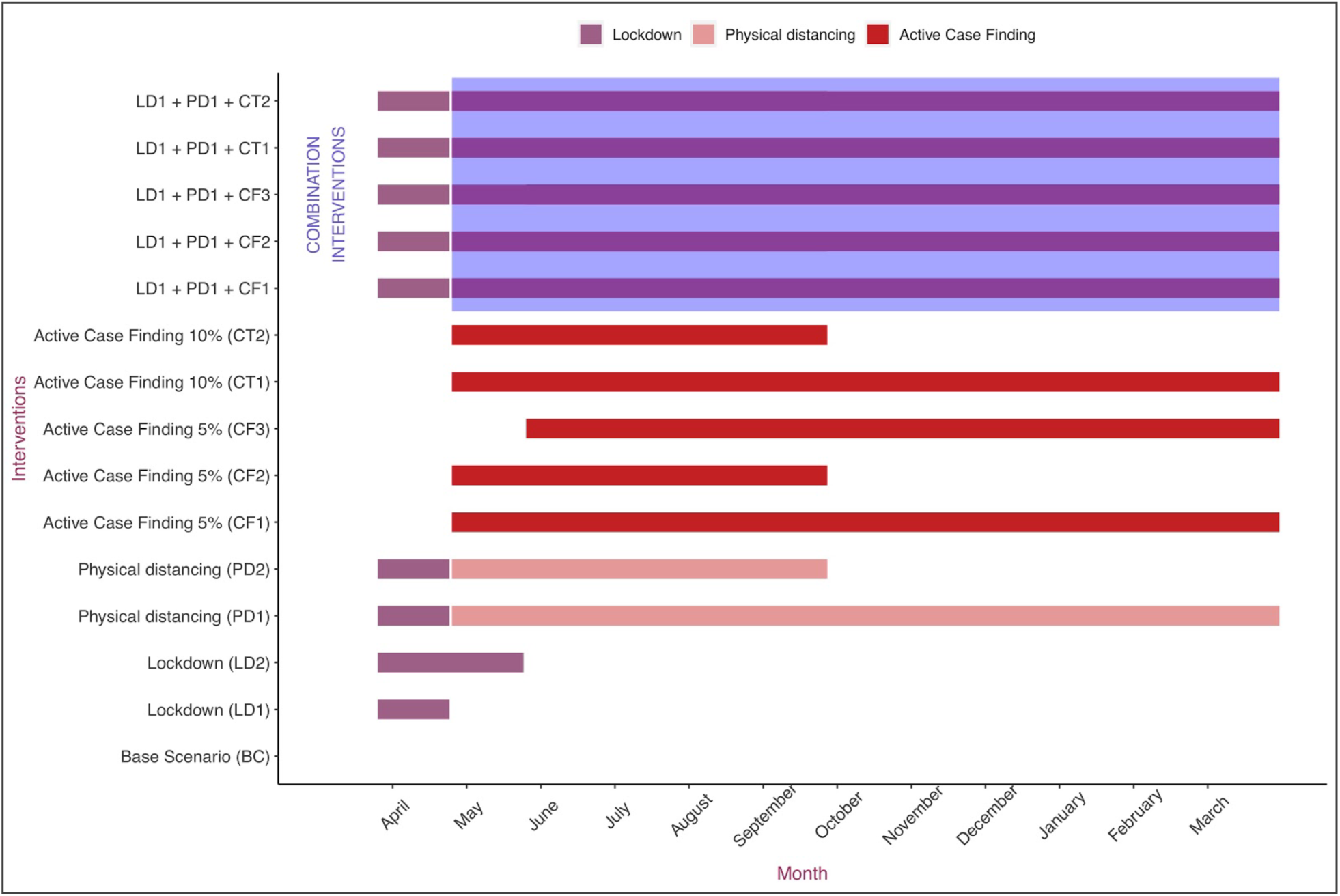
Timeline for the implementation of time-dependent interventions against COVID-19. The y-axis labels represent various interventions that were considered. The colour coded horizontal bars represent the duration the respective intervention is in place. The area on the top shaded in blue represents a combination of interventions. Interventions begin with a lockdown that began on March 24. A one year time duration beginning March 24, 2020 has been considered in this study.

We began our simulations on March 24 when the lockdown began. We assumed that there were 65 exposed and 120 infectious cases--none of which had been detected--on that day. We arrived at these numbers based on the fact that, in an average epidemiological scenario, one additional infectious case of COVID-19 on January 24--when the first case was announced--that went undetected would have led to that many cases by March 24. Our susceptible population was 2·6 million, the total population of the Kathmandu valley. For our analysis, we ignored any additional importation of infections into Kathmandu. First, we simulated a base scenario to estimate the epidemic burden and healthcare demand if no intervention had been instituted. Second, we simulated a scenario where a lockdown is in place for a month starting March 24. We also simulated a lockdown period of 2 months. Third, we simulated a scenario in which a lockdown is in place for a month, followed by physical distancing measures for the entire year, or for half the year. Fourth, we simulated a scenario where enhanced testing, isolation, contact tracing and quarantine is started on April 24, after a month-long lockdown.

Enhanced testing is assumed to identify 5% of the daily burden of infections (exposed and infectious). Finally we tested a strategy combining enhanced testing, isolation, contact tracing and quarantine alongside lockdown and year-long physical distancing measures. We ran our simulations to explore the effects on potential epidemic burden and healthcare demand over a 12 month period. In the supplementary appendix, we also explore the effect of hospital capacity expansion on projected mortality burden.

## Uncertainty of Model Estimates

To explore the uncertainty of our model estimates, we sampled the reproduction number from a uniform distribution in the interval [2, 2·8]. The incubation period was sampled from a uniform distribution in the interval [3, 7] days.

The transmission rate was calculated based on the age and location-specific contact rate. These scenarios are relevant because the burden of undiagnosed cases has been implicated as an important driver of the COVID-19 epidemic. We present the uncertainty of our model estimates in terms of their Inter-Quartile Range (IQR).

## Results

In our hypothetical base scenario of an unmitigated epidemic and no interventions, the epidemic burden is projected to peak at 100 days from March 24. Estimates of epidemic burden for this hypothetical scenario of no intervention are presented in the supplementary appendix (table S1). Demand for general ward hospital beds will peak at 108 days (IQR 97–119) from March 24 and demand for ICU beds peaking at 113 days (IQR 103–124). In this base scenario, peak demand for general ward beds is likely to exceed the current supply of 5400 beds by a factor of 9, and demand for ICU beds is likely to exceed supply by a factor of 25.

These estimates do not account for healthcare demand due to non-COVID-19 illnesses. A 1 month lockdown starting on March 24 will have no effect on the total number of deaths by the end of the year. Demand for healthcare will peak 36 days later (median 144 days, IQR 131–156) as compared to the base scenario, however the number of hospital admissions required will remain the same as the base scenario.

A two-month lockdown will also not make any difference on the number of deaths or the number of hospital admissions required. However, healthcare demand will peak 74 days later as compared to the base scenario (Table 3). Physical distancing measures that reduce overall contact by 35%, introduced at the end of a month long lockdown and in place for a year, will reduce projected deaths by 33% as compared to the base scenario. Demand for healthcare will also fall significantly with peak demand, projected to occur at 245 days (IQR 216–310 days), falling by 65%. ICU demand will fall by 63% and peak a week later. Physical distancing measures that are in place just for the first six months will have a minimal impact in reducing deaths or the peak demand for healthcare, although the peak will occur more than three months later as compared to the base scenario (figures 4 and 5).

**Table 3:**
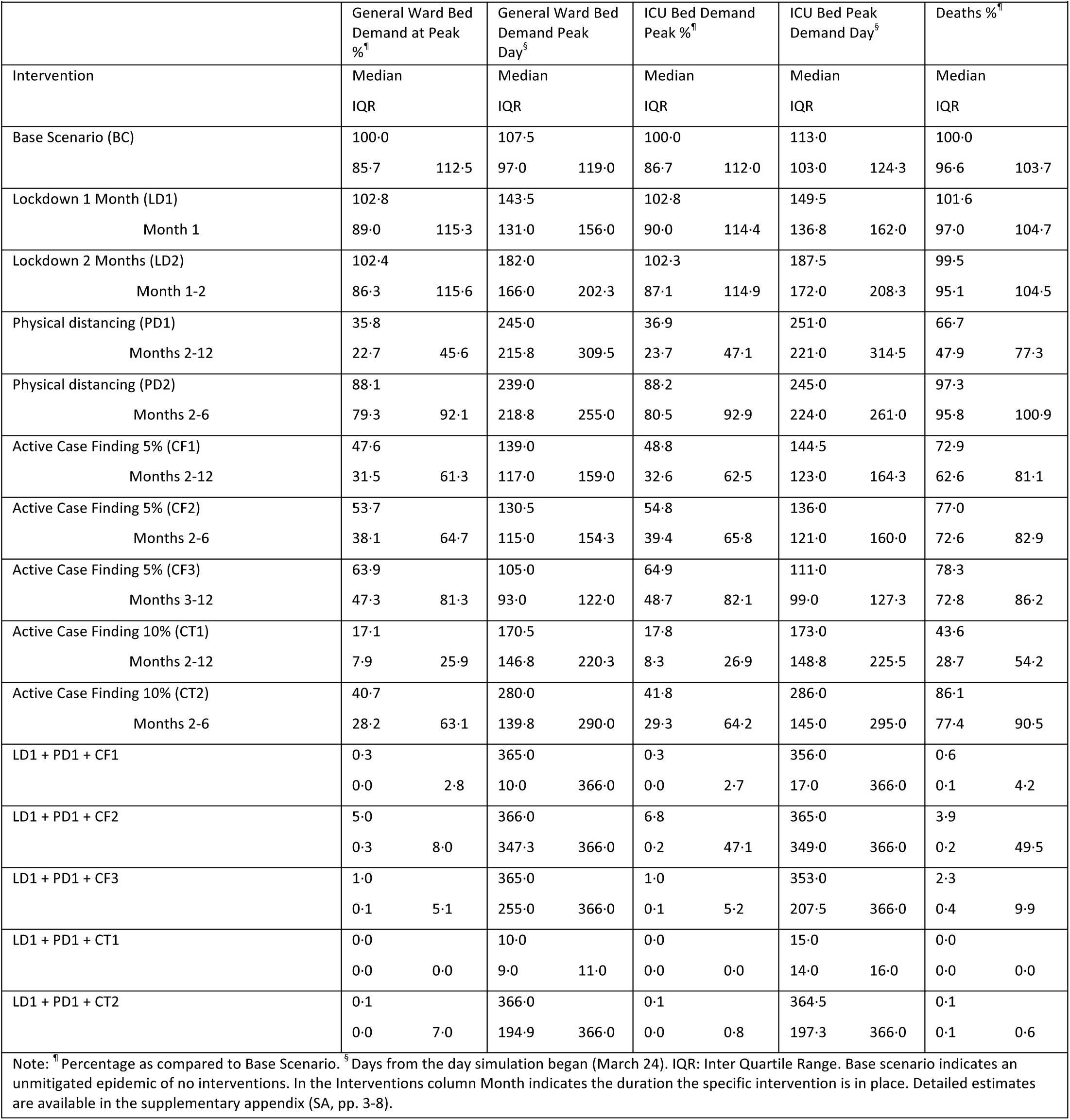
Interventions against COVID-19 and their effectiveness

**Figure 4.**
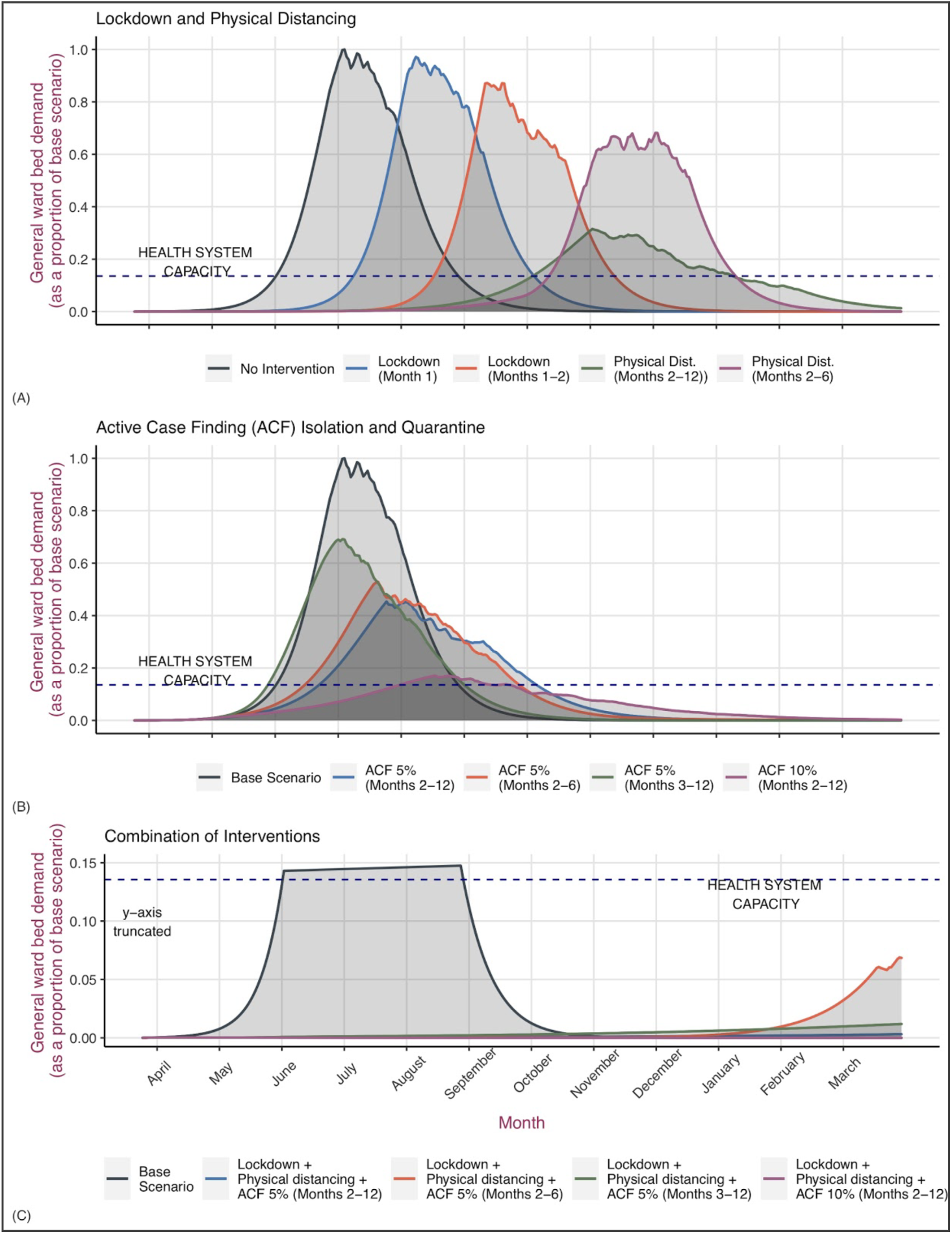
A comparison of the effectiveness of interventions against COVID-19 in reducing the demand for hospital beds. Panel (A) on the top compares lockdown or physical distancing measures implemented for a variable duration as compared to the base scenario of no intervention. Panel (B) in the middle compares active case finding measures with the base scenario. Panel (C) at the bottom compares the effectiveness of a combination of interventions with the base scenario. The blue line represents the health system capacity which is a total of 5400 hospital beds for Kathmandu, including approximately 250 ICU beds. The y-axis has been truncated in Panel (C) to accommodate observations that are closer to the x-axis.

**Figure 5.**
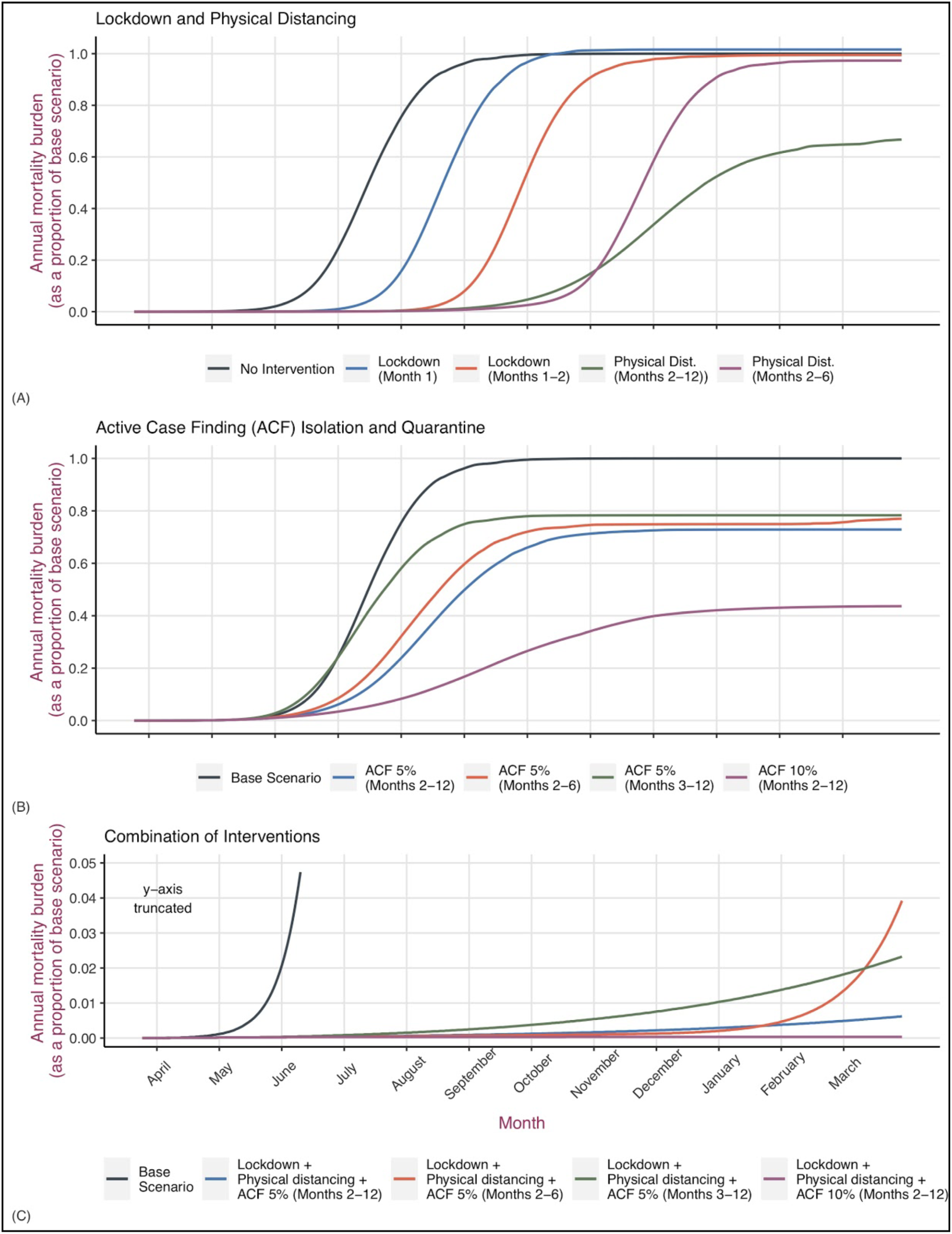
A comparison of the effectiveness of interventions against COVID-19 in reducing mortality burden. Panel (A) on the top compares lockdown or physical distancing measures implemented for a variable duration as compared to the base scenario of no intervention. Panel (B) in the middle compares active case finding measures with the base scenario. Panel (C) at the bottom compares the effectiveness of a combination of interventions with the base scenario of no intervention. The y-axis has been truncated in Panel (C) to accommodate observations that are closer to the x-axis.

Control strategies that are focused on active case finding and isolating infected (exposed and infectious) individuals will lead to greater control of the epidemic burden and significantly reduce the demand for healthcare. If 5% of the prevalent infected people were isolated every day, following a month long lockdown, projected mortality estimates would fall by 27% from the base scenario and demand for healthcare would fall by more than 50%. Healthcare demand would peak at or after day 139 (IQR 117–159 days). If this 5% active case finding intervention were combined with physical distancing measures for the year, total projected deaths by the end of the year would fall by 99.6% with demand for healthcare showing a similar fall. However, if enhanced testing and active case finding intervention were carried out for 6 months and stopped, mortality as well as demand for healthcare increases again. Similarly, starting active case finding at day 60 instead of day 30 would result in 5% less reduction in mortality and 15% less reduction in demand for health services (table 3).

If the active case finding intervention were to be even more enhanced, detecting 10% of cases, but in place for six months and then stopped while physical distancing measures are continued, epidemic burden, demand for healthcare and mortality would continue to remain less than one per cent of the base scenario of an unmitigated epidemic (table 3).

## Discussion

We present a comparison of the effects of a range of possible interventions for preventing and controlling a potential COVID-19 epidemic in a resource limited setting. We find that in the base scenario of an unmitigated epidemic in Kathmandu, the demand for healthcare would significantly exceed supply by a factor of 9 for general ward beds, and by a factor of about 25 for ICU beds. Even with the ongoing lockdown, these outcomes do not change; however a lockdown can prevent an epidemic from escalating and delay its peak, providing vital time to mount other control strategies. Physical distancing interventions that are in place for an entire year would reduce deaths by about a third, and the demand for hospital beds by about two-thirds as compared to the base scenario. The effectiveness of physical distancing measures alone is significantly lower if they are in place for a shorter duration of time. Interventions that aim to actively find and isolate infected individuals are the most effective in reducing the burden of the epidemic, especially when they are combined with other interventions to reduce contact. In the absence of evidence of an uncontrolled epidemic, expansion of hospital capacity is not likely to be the most effective means of reducing potential mortality from an epidemic. Even an expansion of hospital bed capacity by a thousand beds would barely prevent a third of the excess mortality due to the deficit of health services as compared to China (supplementary appendix pp. 2).

Studies and research reviews that evaluate the relative effectivenss of COVID-19 interventions are limited: when they are available they either consider only one type of intervention or have been undertaken in a different context.^8,29,36–38^ Therefore our study is likely to offer several important insights, especially to countries whose context matches the one we considered in this study. Our first insight is on what a lockdown, currently in place in several countries can and can not achieve. While it has bought crucial time to escalate the response against the epidemic by limiting its spread, over the longer run it cannot prevent an unmitigated epidemic if additional interventions are not immediately started. Second, no single intervention will be enough to adequately contain or mitigate the epidemic. Combining a month long lockdown with year long physical distancing measures and enhanced testing and case finding will likely limit the epidemic burden to a minimal. However such synergistic interventions need to begin early and remain in place long enough. If physical distancing as well as active case finding and contact tracing measures are started early enough but stopped in six months, this would still lead to a mortality burden and healthcare demand similar to an unmitigated epidemic. In addition, the more aggressive the early case finding interventions are, the easier long-term COVID-19 control will be.

Third, an enhanced active case finding, isolation, contact tracing intervention appears to be the cornerstone of any successful control strategy. This is understandable given recent findings that almost half of the transmission may originate from pre-symptomatic individuals.^39^ Fourth, in the face of an unmitigated epidemic, health systems in resource limited settings are likely to be significantly overwhelmed, resulting in mortality rates that could be double the expected rates. This is despite accounting for the younger median age of the population. As our results demonstrate, once an unmitigated epidemic is underway, even a significant expansion of hospital bed capacity will not be enough to adequately curtail the excess mortality burden. Hence a control strategy that focuses more on the expansion of hospital capacity even when combined with lockdowns and physical distancing measures, without enhanced testing and active case finding is likely to lead to significant epidemic burden and deaths.

Our analysis using time controlled interventions allows us to evaluate the type and duration of interventions required to mount an effective control strategy. A month long lockdown and physical distancing interventions combined with an active case finding intervention instituted early is likely to effectively control a potential epidemic, however physical distancing and testing interventions have to continue for the year.

Assuming that there has been no additional importation of SARS-Cov2 infection beyond the one case we considered, given the lockdown, a very conservative estimated burden of infections in Kathmandu may be less than two hundred. Even then, assuming a 0.4–0.5% yield, daily tests would need to increase to about 2000–2500 in Kathmandu to actively find 5% of the current infection burden. Testing volumes however, remain a fraction of that. Only less than 10 000 RT-PCR based tests have been carried out in the entire country as of April 22. A similar number of antibody based rapid diagnostic tests have been carried out as well, but their accuracy has been suspect. Inaccuracy of currently available tests has made case finding difficult.^40^

A control strategy that targets finding a fixed percentage of the daily burden of infections scales in a geometric fashion making it programmatically challenging to implement in places with limited capacity for effective public health interventions. However, even while many countries with limited resources are finding it difficult to sufficiently escalate testing, isolation and contact tracing, some others have successfully done so.^41^ Community and local government led best practices are beginning to emerge where local governments and communities are facilitating testing and quarantine of possible contacts.^11,42,43^ Technology based solutions may additionally help in contact-tracing and following up in individuals in isolation and quarantine.^44^

Given the current focus on a lockdown, we have not modelled the effect of targeted physical distancing measures like school closures or measures that target the elderly. However, these measures could be significantly important, given people in Nepal tend to live in intergenerational households that have significant contact between family members. While our use of a Nepal specific synthetic contact matrix minimises some of these concerns, the effect of targeted physical distancing measures needs to be evaluated in their local context. This could be an important area for further work.

Our analysis makes significant simplifying assumptions. First, we assume that there has been no additional importation of COVID-19 cases in Kathmandu. This assumption is not true, since there have been additional imported cases diagnosed since the first case, however this does not invalidate our findings. We assume that everyone is equally susceptible to the virus and that there is no immunity. However, as yet it is unclear if this assumption holds true. We make assumptions about the effectiveness of a lockdown and home isolation. We also assume that it would be possible to enforce physical distancing measures that reduce contact between individuals by about a third for the entire year, an assumption that might not hold over time as the public grows weary of such measures.^45^ In addition, prolonged physical distancing measures could themselves lead to adverse health, economic and well-being outcomes--an issue that is likely to severely impact many countries that do not have adequate social safety provisions. This is another area for future work. Our use of a compartmental model offers a computationally simpler method to model the epidemic, although this approach is not likely to be methodologically valid if an epidemic is yet to be established.^46,47^ We overcame this limitation by assuming that there already were about 185 infected individuals and that local transmission was under way when we began our model simulation.

Our study offers important insights on mounting an effective response against the COVID-19 epidemic in a resource limited setting. As we have outlined above, an unmitigated COVID-19 epidemic has the potential to cause significant mortality that will be exacerbated by the unmet demand for health services. Our findings suggest that the best control strategy against the epidemic is a combination of interventions that aim to identify and isolate infected individuals and reduce contact between individuals by means of ongoing physical distancing measures. A lockdown can prevent the escalation of the epidemic, but is likely to be of limited value if no additional control measures are put in place.

## Data Availability

All data used in this study have been referenced in the manuscript are available online.

## Declarations

### Author Contributions

KRP designed the study, obtained the data, built the model, conducted the analysis and wrote the manuscript. AS designed the study, obtained the data, interpreted the results and edited the manuscript. BKh reviewed the model and the analysis and edited the manuscript. BKo reviewed the analysis and edited the manuscript.

### Ethics Committee Approval

No ethical approval was sought for this study beause this modeling study does not use any patient identifying information and no primay data was collected during this study.

## Bibliography

1 WHO. Coronavirus (COVID-19) events as they happen. Rolling updates on coronavirus disease. 2020; published online April 20. https://www.who.int/emergencies/diseases/novel-coronavirus-2019/events-as-they-happen (accessed April 23, 2020).

2 World Health Organization. Coronavirus disease (COVID-2019) situation reports. 2020; published online April 21. https://www.who.int/emergencies/diseases/novel-coronavirus-2019/situation-reports (accessed April 22, 2020).

3 Nkengasong JN, Mankoula W. Looming threat of COVID-19 infection in Africa: act collectively, and fast. Lancet 2020; 395: 841–2.

4 Makoni M. Africa prepares for coronavirus. Lancet 2020; 395: 483.

5 Schafer H. COVID-19 will hit South Asia hard. We are fighting back. World Bank Blogs. 2020; published online April 2. https://blogs.worldbank.org/endpovertyinsouthasia/covid-19-will-hit-south-asia-hard-we-are-fighting-back (accessed April 23, 2020).

6 WHO. COVID 19 strategy update. 2020; published online April 14.

7 Critical preparedness, readiness and response actions for COVID-19. https://www.who.int/emergencies/diseases/novel-coronavirus-2019/technical-guidance/critical-preparedness-readiness-and-response-actions-for-covid-19 (accessed April 22, 2020).

8 Hellewell J, Abbott S, Gimma A, et al. Feasibility of controlling COVID-19 outbreaks by isolation of cases and contacts. Lancet Glob Health 2020; 8: e488–96.

9 Bedford J, Enria D, Giesecke J, et al. COVID-19: towards controlling of a pandemic. Lancet 2020; 395: 1015–8.

10 Wang CJ, Ng CY, Brook RH. Response to COVID-19 in Taiwan: Big Data Analytics, New Technology, and Proactive Testing. JAMA 2020; published online March 3. DOI:10.1001/jama.2020.3151.

11 Nyugen HK. Vietnam’s Low-Cost COVID-19 Strategy by Hong Kong Nguyen. Project Syndicate. 2020; published online April 8. https://www.project-syndicate.org/commentary/vietnam-low-cost-success-against-covid19-by-hong-kong-nguyen-2020-04 (accessed April 22, 2020).

12 Vu M, Tran BT. The Secret to Vietnam’s COVID-19 Response Success. The Diplomat. 2020; published online April 18. https://thediplomat.com/2020/04/the-secret-to-vietnams-covid-19-response-success/ (accessed April 22, 2020).

13 Chung D. Korea’s response to COVID-19: Early lessons in tackling the pandemic. https://blogs.worldbank.org/eastasiapacific/koreas-response-covid-19-early-lessons-tackling-pandemic (accessed April 22, 2020).

14 Zastrow M. South Korea is reporting intimate details of COVID-19 cases: has it helped? Nature 2020; published online March 18. DOI:10.1038/d41586-020-00740-y.

15 Wu Z, McGoogan JM. Characteristics of and Important Lessons From the Coronavirus Disease 2019 (COVID-19) Outbreak in China: Summary of a Report of 72 314 Cases From the Chinese Center for Disease Control and Prevention. JAMA 2020; published online Feb 24. DOI:10.1001/jama.2020.2648.

16 Chen S, Yang J, Yang W, Wang C, Bärnighausen T. COVID-19 control in China during mass population movements at New Year. Lancet 2020; 395: 764–6.

17 HEOC. COVID-19 Nepal situation report no. 69. 2020; published online April 18.

18 Pradhan TR. Nepal goes under lockdown for a week starting 6am Tuesday. The Kathmandu Post. 2020; published online March 23. https://kathmandupost.com/national/2020/03/23/nepal-goes-under-lockdown-for-a-week-starting-6am-tuesday (accessed April 22, 2020).

19 Brauer F. Compartmental models in epidemiology. In: Brauer F, van den Driessche P, Wu J, eds. Mathematical Epidemiology. Berlin, Heidelberg: Springer Berlin Heidelberg, 2008: 19–79.

20 Del Valle SY, Hyman JM, Chitnis N. Mathematical models of contact patterns between age groups for predicting the spread of infectious diseases. Math Biosci Eng 2013; 10: 1475–97.

21 Prem K, Cook AR, Jit M. Projecting social contact matrices in 152 countries using contact surveys and demographic data. PLoS Comput Biol 2017; 13: e1005697.

22 Ferretti L, Wymant C, Kendall M, et al. Quantifying SARS-CoV-2 transmission suggests epidemic control with digital contact tracing. Science 2020; published online March 31. DOI:10.1126/science.abb6936.

23 Li Q, Guan X, Wu P, et al. Early Transmission Dynamics in Wuhan, China, of Novel Coronavirus-Infected Pneumonia. N Engl J Med 2020; 382: 1199–207.

24 Kucharski AJ, Russell TW, Diamond C, et al. Early dynamics of transmission and control of COVID-19: a mathematical modelling study. Lancet Infect Dis 2020; published online March 11. DOI:10.1016/S1473-3099(20)30144-4.

25 Woelfel R, Corman VM, Guggemos W, et al. Clinical presentation and virological assessment of hospitalized cases of coronavirus disease 2019 in a travel-associated transmission cluster. *medRxiv* 2020; published online March 8. DOI:10.1101/2020.03.05.20030502.

26 Verity R, Okell LC, Dorigatti I, et al. Estimates of the severity of coronavirus disease 2019: a model-based analysis. Lancet Infect Dis 2020; published online March 30. DOI:10.1016/S1473-3099(20)30243-7.

27 Guan W-J, Ni Z-Y, Hu Y, et al. Clinical characteristics of coronavirus disease 2019 in China. N Engl J Med 2020; published online Feb 28. DOI:10.1056/NEJMoa2002032.

28 Paneru H. Intensive care units in the context of COVID-19 in Nepal: current status and need of the hour. JSAN 2020; 7.

29 Prem K, Liu Y, Russell TW, et al. The effect of control strategies to reduce social mixing on outcomes of the COVID-19 epidemic in Wuhan, China: a modelling study. Lancet Public Health 2020; published online March 25. DOI:10.1016/S2468-2667(20)30073-6.

30 Zhou F, Yu T, Du R, et al. Clinical course and risk factors for mortality of adult inpatients with COVID-19 in Wuhan, China: a retrospective cohort study. Lancet 2020; 395: 1054–62.

31 Saunders-Hastings P, Quinn Hayes B, Smith R, Krewski D. Modelling community-control strategies to protect hospital resources during an influenza pandemic in Ottawa, Canada. PLoS ONE 2017; 12: e0179315.

32 Coburn BJ, Wagner BG, Blower S. Modeling influenza epidemics and pandemics: insights into the future of swine flu (H1N1). BMC Med 2009; 7: 30.

33 Halloran ME, Ferguson NM, Eubank S, et al. Modeling targeted layered containment of an influenza pandemic in the United States. Proc Natl Acad Sci USA 2008; 105: 4639–44.

34 Ferguson NM, Cummings DAT, Fraser C, Cajka JC, Cooley PC, Burke DS. Strategies for mitigating an influenza pandemic. Nature 2006; 442: 448–52.

35 Longini IM, Nizam A, Xu S, et al. Containing pandemic influenza at the source. Science 2005; 309: 1083–7.

36 Nussbaumer-Streit B, Mayr V, Dobrescu AI, et al. Quarantine alone or in combination with other public health measures to control COVID-19: a rapid review. Cochrane Database Syst Rev 2020; 4: CD013574.

37 Koo JR, Cook AR, Park M, et al. Interventions to mitigate early spread of SARS-CoV-2 in Singapore: a modelling study. Lancet Infect Dis 2020; published online March 23. DOI:10.1016/S1473-3099(20)30162-6.

38 Singh R, Adhikari R. Age-structured impact of social distancing on the COVID-19 epidemic in India. *arXiv* 2020; published online March 26.

39 He X, Lau EHY, Wu P, et al. Temporal dynamics in viral shedding and transmissibility of COVID-19. Nat Med 2020; published online April 15. DOI:10.1038/s41591-020-0869-5.

40 New Spotlight. Nepal Bars Using Rapid Test Kits Recently Imported From China. New Spotlight. 2020; published online April 2. https://www.spotlightnepal.com/2020/04/02/nepal-bars-using-rapid-test-kits-recently-imported-china/ (accessed April 23, 2020).

41 Worldometers. Reported Cases and Deaths by Country, Territory, or Conveyance. Worldometers. info. 2020; published online April 22. https://www.worldometers.info/coronavirus/#countries (accessed April 22, 2020).

42 Biswas S. Coronavirus: How India’s Kerala state “flattened the curve” - BBC News. BBC News. 2020; published online April 16. https://www.bbc.com/news/world-asia-india-52283748 (accessed April 17, 2020).

43 Corona Virus: Status Update of Sudur Paschim Province—Inseconline. http://inseconline.org/en/news/corona-virus-situation-of-far-west-province/ (accessed April 23, 2020).

44 Greenberg A. How Apple and Google Are Enabling Covid-19 Bluetooth Contact-Tracing. Wired. 2020; published online April 10. https://www.wired.com/story/apple-google-bluetooth-contact-tracing-covid-19/ (accessed April 22, 23 2020).

45 Parmet WE, Sinha MS. Covid-19 - The Law and Limits of Quarantine. N Engl J Med 2020; 382: e28.

46 Roberts M, Andreasen V, Lloyd A, Pellis L. Nine challenges for deterministic epidemic models. Epidemics 2015; 10: 49–53.

47 Blackwood JC, Childs LM. An introduction to compartmental modeling for the budding infectious disease modeler. Letters in Biomathematics 2018; 5: 195–221.

